# Psychological morbidities and fatigue in patients with confirmed COVID-19 during disease outbreak: prevalence and associated biopsychosocial risk factors

**DOI:** 10.1101/2020.05.08.20031666

**Authors:** Rongfeng Qi, Wei Chen, Saiduo Liu, Paul M. Thompson, Long Jiang Zhang, Fei Xia, Fang Cheng, Ailing Hong, Wesley Surento, Song Luo, Zhi Yuan Sun, Chang Sheng Zhou, Lingjiang Li, Xiangao Jiang, Guang Ming Lu

## Abstract

**Objective:** The coronavirus disease 2019 (COVID-19) – a novel and highly infectious pneumonia – has now spread across China and beyond for over four months. However, its psychological impact on patients is unclear. We aim to examine the prevalence and associated risk factors for psychological morbidities and fatigue in patients with confirmed COVID-19 infection.

**Methods:** Amidst the disease outbreak, 41 out of 105 COVID-19 patients in a local designated hospital in China were successfully assessed using a constellation of psychometric questionnaires to determine their psychological morbidities and fatigue. Several potential biopsychosocial risk factors (including pre-existing disabilities, CT severity score of pneumonia, social support, coping strategies) were assessed through multivariable logistic regression analyses to clarify their association with mental health in patients.

**Results:** 43.9% of 41 patients presented with impaired general mental health, 12.2% had post-traumatic stress disorder (PTSD) symptoms, 26.8% had anxiety and/or depression symptoms, and 53.6% had fatigue. We did not find any association between pneumonia severity and psychological morbidities or fatigue in COVID-19 patients. However, high perceived stigmatization was associated with an increased risk of impaired general mental health and high perceived social support was associated with decreased risk. Besides, negative coping inclination was associated with an increased risk of PTSD symptoms; high perceived social support was associated with a decreased risk of anxiety and/or depression symptoms.

**Conclusions:** Psychological morbidities and chronic fatigue are common among COVID-19 patients. Negative coping inclination and being stigmatized are primary risk factors while perceived social support is the main protective factor.

## INTRODUCTION

In December 2019, the novel coronavirus disease 2019 (COVID-19) outbreak occurred in Wuhan, Hubei Province, China^1,2^. It has quickly spread across China and beyond, resulting in total confirmed cases 3,646,103 and 252,407 confirmed deaths across the world, as of May 5, 2020, according to Worldometers (https://www.worldometers.info/coronavirus/). To lower the risk of further disease transmission, several methods have been urgently implemented in many countries, such as drastic limitations on public transport, early identification followed by isolation of suspected and diagnosed cases, along with establishment of new isolation units and even hospitals^3^.

At present, most energy and resources tend to be directed towards physical well-being in confirmed and suspected cases of COVID-19, but psychological morbidities in patients are neglected and have yet to be formally evaluated^4^. The National Health Commission of China released a notification of basic principles for emergency psychological crisis interventions for the COVID-19 on January 26, 2020, and later, guidelines for psychological assistance hotlines dealing with the COVID-19 epidemic on February 2, 2020. On March 11, the World Health Organization (WHO) declared COVID-19 a worldwide pandemic, as it has rapidly spread to more than 150 countries and regions. There is still much work to be done to increase awareness and respond to the psychological impact of this novel and highly infectious pneumonia^3,4^. As with prior healthcare crises such as the 2003 severe acute respiratory syndrome (SARS)^5^ and Middle East respiratory syndrome epidemic (MERS)^6,7^, the emerging COVID-19 outbreak is expected to result in immediate and even long-term mental health problems. During and after the SARS and MERS outbreaks, infected patients were commonly reported to experience psychological distress, anxiety or depression symptoms, psychiatric disorders, and chronic fatigue^6–10^. Several recent studies have reported that the front-line healthcare workers are vulnerable to the emotional impact of the coronavirus^11,12^, however, little is known about the effects of the coronavirus on patients with laboratory-confirmed COVID-19 infection and associated risk factors.

In this study, we aim to characterize psychological morbidities and fatigue in patients confirmed to have COVID-19 infection amidst the disease outbreak. We hypothesized that psychological morbidities and fatigue would be common in COVID-19 patients. Furthermore, a biopsychosocial model – which accounts for pre-existing disabilities, history of psychiatric disorders, social support, coping strategies, and personality traits – may explain the development of psychological problems in individuals affected by healthcare crises such as SARS and MERS^7^. In this study, we collected information on several biopsychosocial risk factors to clarify their relationship with mental health in COVID-19 patients. We further hypothesized that specific biopsychosocial factors – such as social support, which was commonly reported to have an association with SARS-related psychological problems^13^ – may be associated with COVID-19-related psychological morbidities and fatigue.

## SUBJECTS and METHODS

### Subjects

This cross-sectional study on psychological morbidities and fatigue in COVID-19 patients was conducted within a local designated hospital in February 2020. Amidst the COVID-19 outbreak, 105 patients with laboratory-confirmed COVID-19 infection had received treatment in the isolation wards of this designated hospital in China. This study was approved by Medical Research Ethics Committee of Jinling Hospital; written informed consent was obtained from all participants. Among these patients, those who are classified as non-severe types of COVID-19, not illiterate, and without any major neurological conditions were invited to participate in the study. An earlier notification regarding the study was conveyed to all eligible patients in the form of personalized letters, a few days before the study began. Subsequently, self-administered questionnaires were distributed to patients who agreed to participate in this study via the respective isolation ward managers, who also collected the responses. All contents and results recorded on paper documents that were taken into the isolation wards were transmitted out as camera images.

### Measures

Each patient who agreed to participate in the study was assessed using a set of self-administered questionnaires, which included the General Health Questionnaire-12 (GHQ-12)^14^, PTSD CheckList-Civilian Version (PCL-C)^15^, Zung Self-Rating Anxiety Scale (SAS)^16^, Zung Self-Rating Depression Scale (SDS)^17^, Fatigue Scale-14 (FS-14)^18^, Chinese Social Support Rating Scale (SSRS)^19^, and individual Simple Coping Style Questionnaire (SCSQ)^20^. The degree of perceived social stigmatization was also assessed by asking participants to rate whether they perceived any stigmatization due to their infection status, on a scale of 1 (no stigmatization) to 4 (always perceived stigmatization)^10^.

The GHQ-12 items are widely used for screening general mental health in the community, which includes 12 items on a 4-point response scale, and is scored in a bimodal fashion: symptom presentation: ‘‘not at all’’ (0); ‘‘same as usual’’ (0); ‘‘rather more than usual’’ (1); and ‘‘much more than usual’’ (1). The cutoff score for the total score was set at 3^21^.

The PCL-C is a 17-item self-report measure reflecting DSM-IV symptoms of PTSD with a validated cutoff of 50^15^.

The cutoff values for SAS and SDS standardized scores are set at 50 and 53^16,17^, respectively; and set at 4 for fatigue^18^.

The SSRS contains three subscales of social support: ① subjective or perceived support, which refers to an individual’s perceptions of the interpersonal network that he or she can count on; ② objective support, which reflects the actual support that an individual received; and ③ the utility of support, which indicates the pattern of support-seeking behavior. Higher scores indicate stronger corresponding social support.

The SCSQ contains measurements for both active and negative coping. The scale of each SCSQ item uses a 4-level Likert score standard, graded from 3 (stands for regular use) to 0 (no use). The scores for active and negative coping are calculated independently; a higher score suggests the inclination to adopt the corresponding coping style.

In addition, demographic data that included age, sex, educational level, marital status, history of psychiatric disorders, pre-existing health conditions (including cardiovascular disease, diabetes, hypertension, chronic obstructive pulmonary disease, chronic liver and kidney diseases, and malignancy), and clinical symptoms were extracted from inpatient medical records. Besides, the time intervals between initial symptom onset, hospitalization date, most recent computed tomography (CT) scan and psychometric assessments were calculated for each patient. The CT severity score^22^ of pneumonia was assessed individually as follows: each lung was defined as having 10 segments (both upper lobes, *n*=3, middle lobe and the lingular division, *n*=2, both lower lobes, *n*=5). The score for each segment was recorded: 0 for normal and 1 for presence of any lesion, regardless of opacity and extent. The maximum theoretic score was 20 points if all segments are involved. CT severity score is expressed as (*n*)/20 × 100%, where *n* is the number of involved segments.

### Statistical analysis

Patients were classified into 2 groups based on each of 4 main outcome measures – general mental health, PTSD symptoms, anxiety and/or depression symptoms, and chronic fatigue problems. SPSS version 25 (IBM Corp, Armonk, New York, USA) was used to analyze the clinical and psychological data. The normality of the quantitative data was checked using a Kolmogorov-Smirnov 1-sample test. The group comparisons on continuous variables were assessed by *t* test for normally distributed data and Mann–Whitney U test for non-normally distributed data, while categorical variables were analyzed by Pearson χ^2^ test or Fisher’s exact test where appropriate. Multivariable logistic regression analyses were conducted to determine biopsychosocial factors (sex, age, educational level, marital status, history of psychiatric disorders, pre-existing disabilities, clinical symptoms, CT severity score, social support scales, coping strategies, and perceived stigmatization) that were significantly associated with psychological morbidities and fatigue. The cutoff point for the selected variables for multivariable logistic regression was fixed at *P* ≤ 0.10 ^10^. The level of statistical significance for group comparisons and logistic regression analyses was set to *P*=0.05 (two-sided).

## RESULTS

### Flowchart of the study population

The flowchart of the study population is shown in **Figure 1**. There were a total of 105 confirmed COVID-19 patients who had received treatment in the isolation wards of a local designated hospital in China. In contrast to the entire population of patients within the largest study of COVID-19^2^ to date (1,099 patients from 552 hospitals in 31 provinces/provincial municipalities in China), our study participants showed no significant differences in terms of sex ratio and age (to compare the whole population in that study to our cluster of participants, respectively: percentage of females, 41.8% vs 44.8% [*P* =0.56]; median age, 47 years vs 46 years [*P* = 0.08]); but our cohort had marginally lower disease severity ratio (participants diagnosed with severe type, 15.7% vs 8.57% [*P* =0.05]).

**Figure 1.**
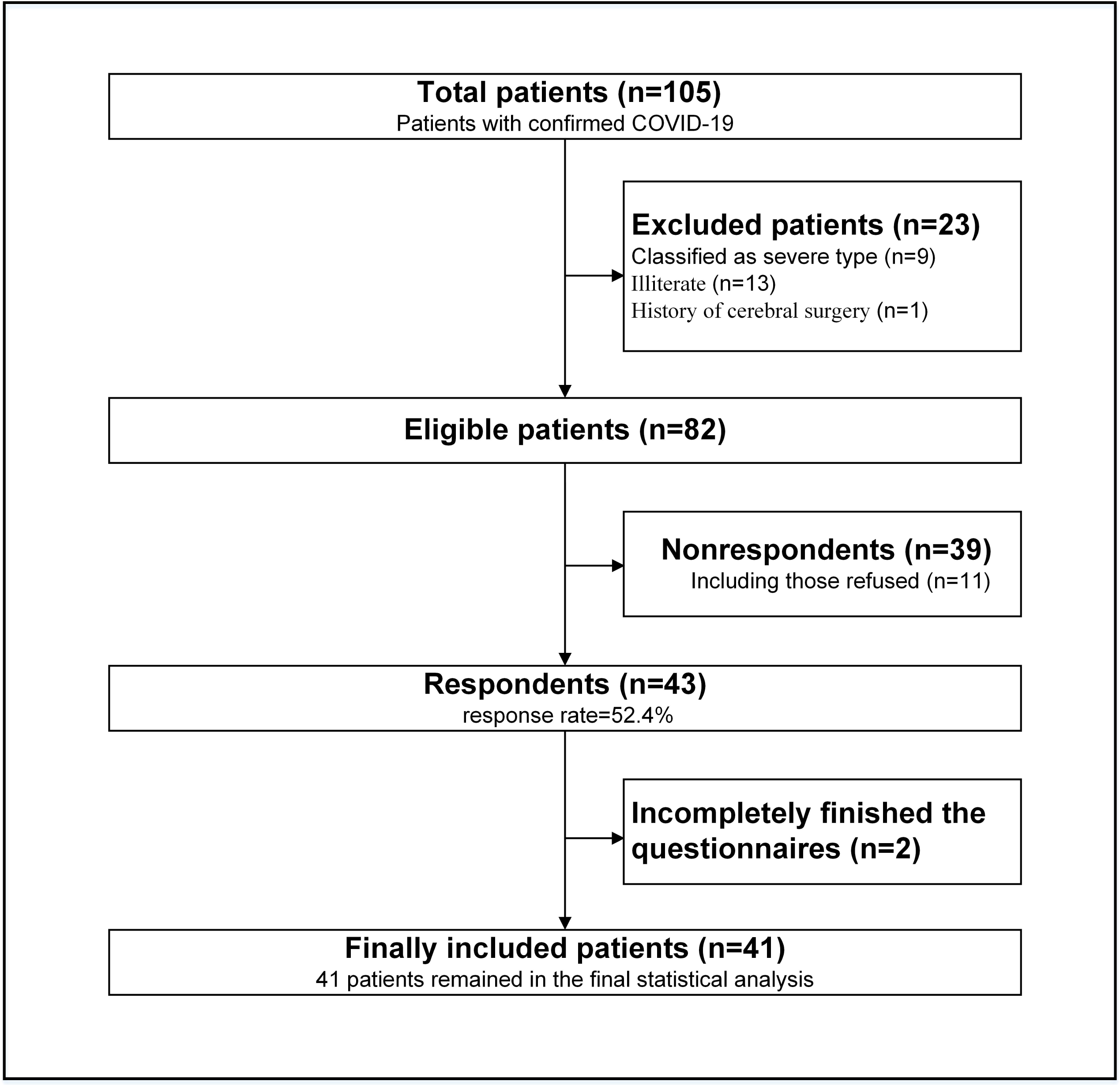
The flowchart of the study population in this study. COVID-19 refers to the coronavirus disease 2019.

In our study, we excluded patients who were severely ill (classified as severe or critically severe types) (n=9); those who were illiterate (n=13); and those with a current or history of major neurological conditions (1 patient was excluded for having a history of surgery for intracranial aneurysm). The remaining 82 patients were invited to participate. 43 patients (response rate = 52.4%) agreed to participate in the study, and were then asked to complete a set of questionnaires.

The mean (SD) age of the respondents was 40.1 (10.1) years; 58.1% of the individuals were female. Most were married (88.4%). Compared to nonrespondents, respondents were more likely to be female, but the 2 groups did not differ in age, educational level, pre-existing medical comorbidities, CT severity score, clinical symptoms, or steroid therapy status (**Table 1**). None of the respondents or nonrespondents have been admitted to intensive care unit (ICU), or reported with a history of psychiatric disorders.

**Table 1:**
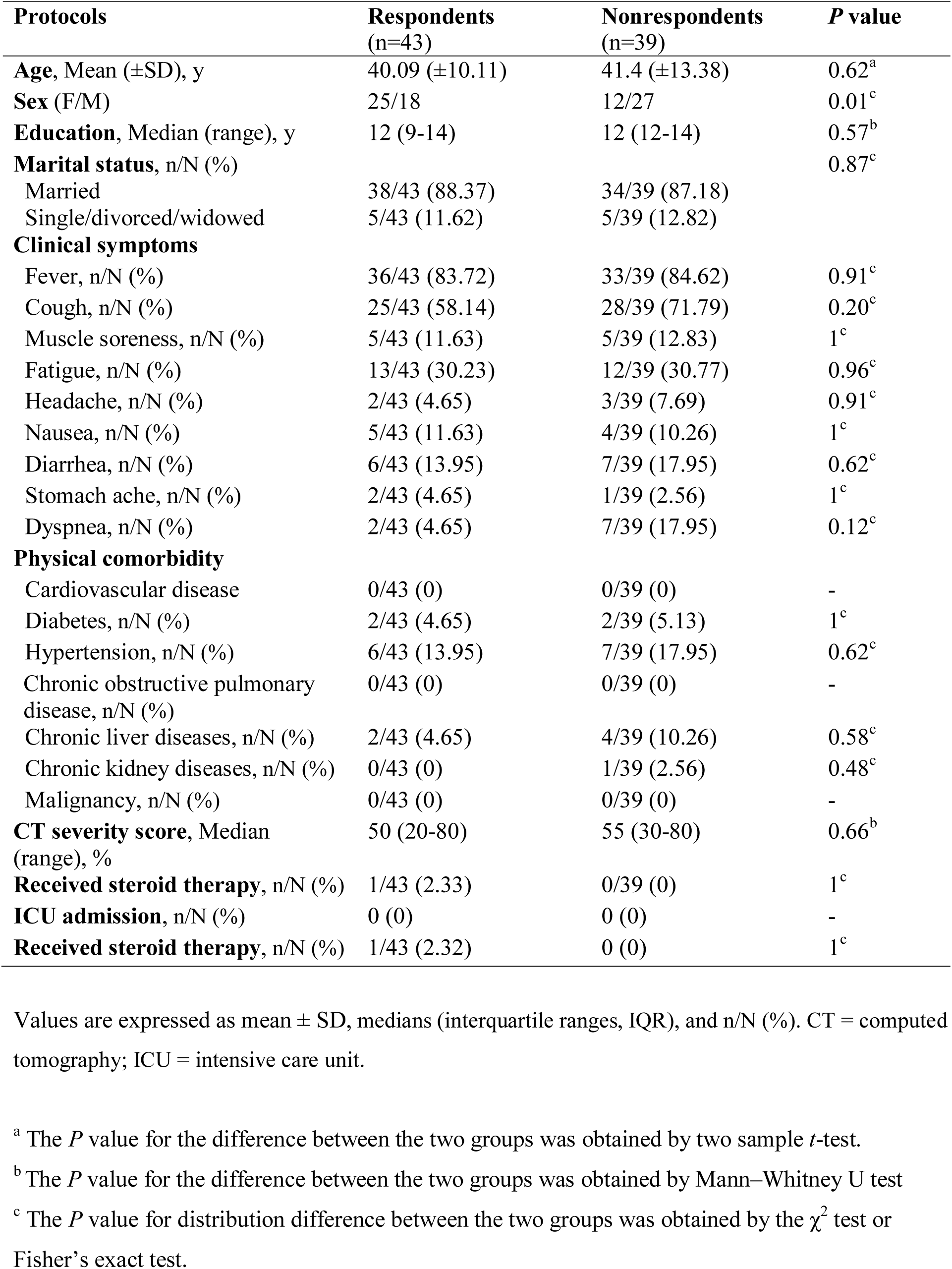
Comparisons of respondents to nonrespondents in this study with confirmed COVID-19.

Two respondents were further excluded as they finished less than 1/3 of all the questionnaires, and were unwilling to complete them. For the remaining 41 participants, the median time interval between symptom onset and psychometric assessment was 27 days (interquartile range: 23–28), between hospitalization and psychometric assessment was 27 days (22–28), and between the latest CT scan and psychometric assessment was 21 days (11–24).

### General mental health in COVID-19 patients

18 out of 41 patients (43.9%) presented with general mental health problems. There were no significant differences between patients with and without general mental health problems in terms of age, educational level, marital status, coping style, stigmatization, physical comorbidities, CT severity score, clinical symptoms, or any intervals from initial symptom onset, hospitalization date, and most recent CT scan to psychometric assessment (**Table 2**). However, patients with general mental health problems were more likely to be female, had higher perceived stigmatization but lower objective and subjective/perceived social support scores. Results of logistic regression analyses showed that having high perceived stigmatization (odds ratio [OR], 3.29; 95% confidence interval [CI], 1.18–9.17; *P* = 0.02) was associated with a greater risk of general mental health problems, while having a high perceived support score (OR, 0.78; 95% CI, 0.62–0.98; *P* = 0.04) was associated with a lower risk (**Table 3**).

**Table 2.**
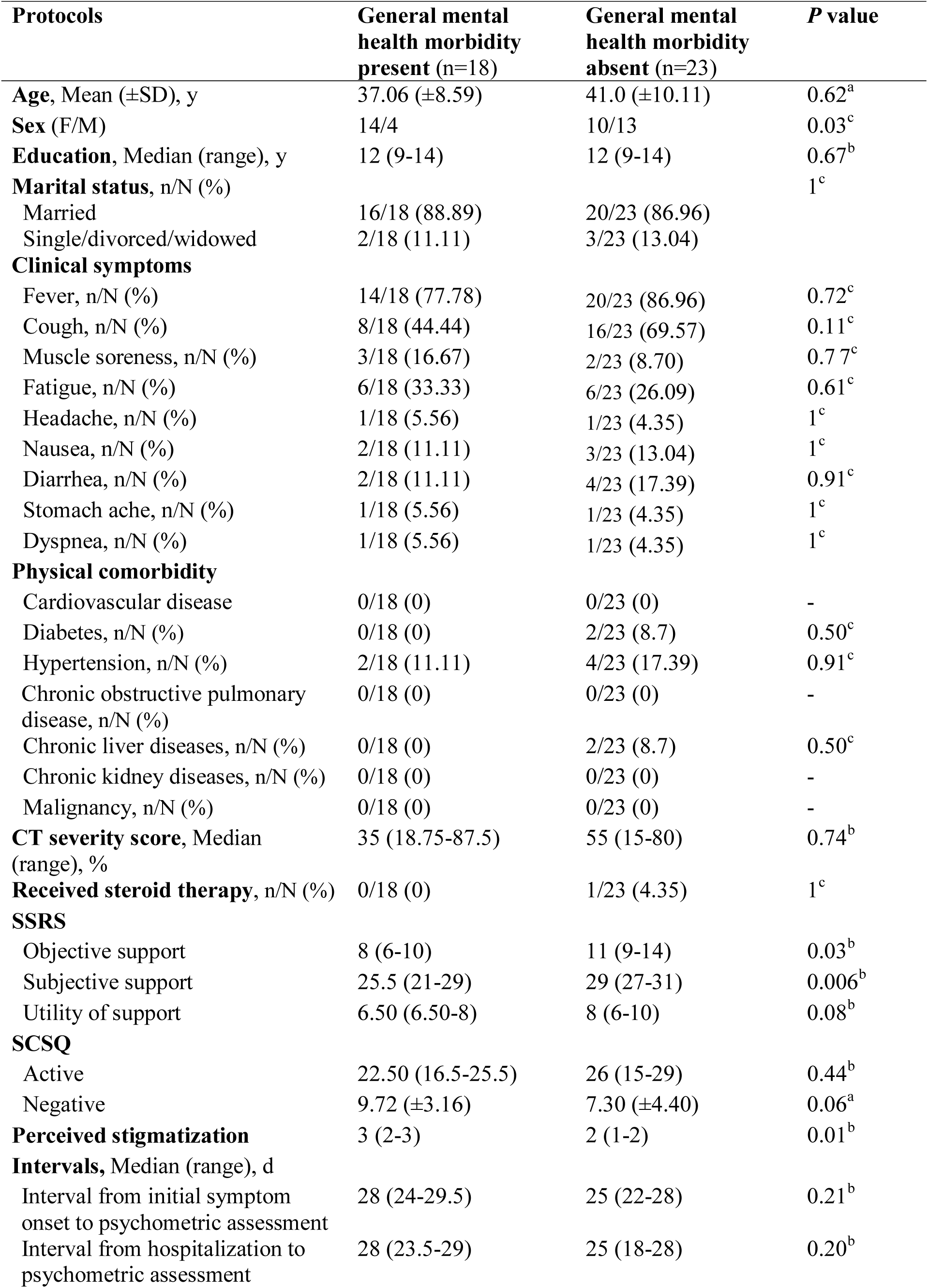
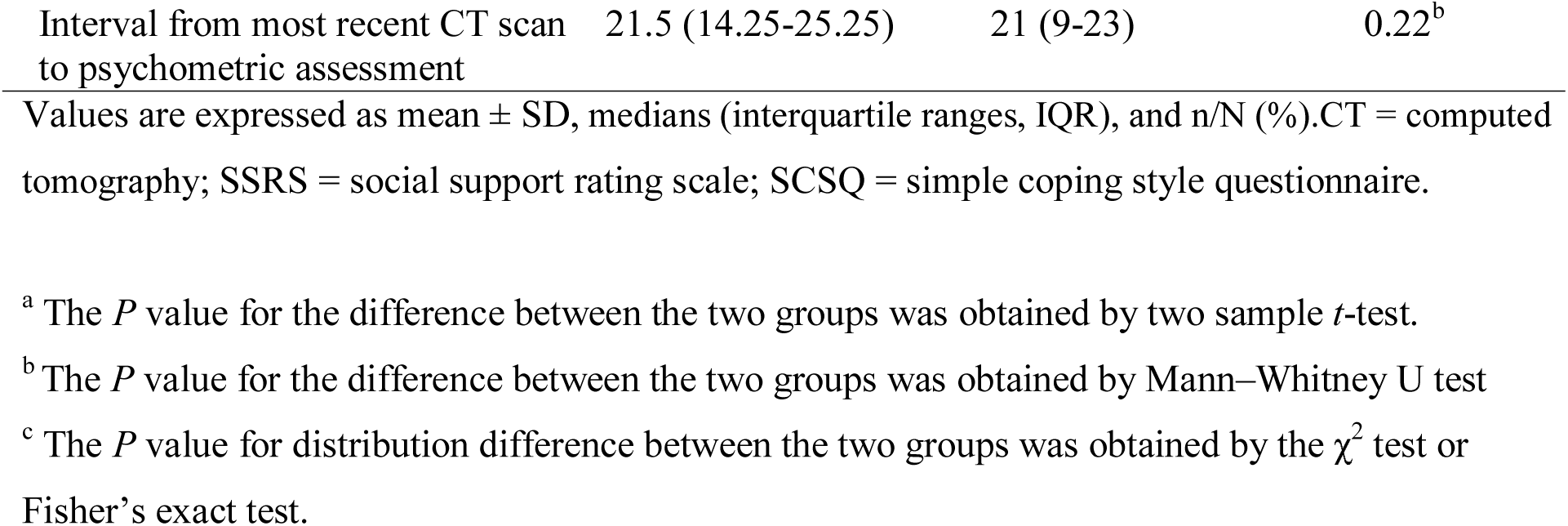
Comparisons of COVID-19 patients with and without general mental health problems.

**Table 3.**
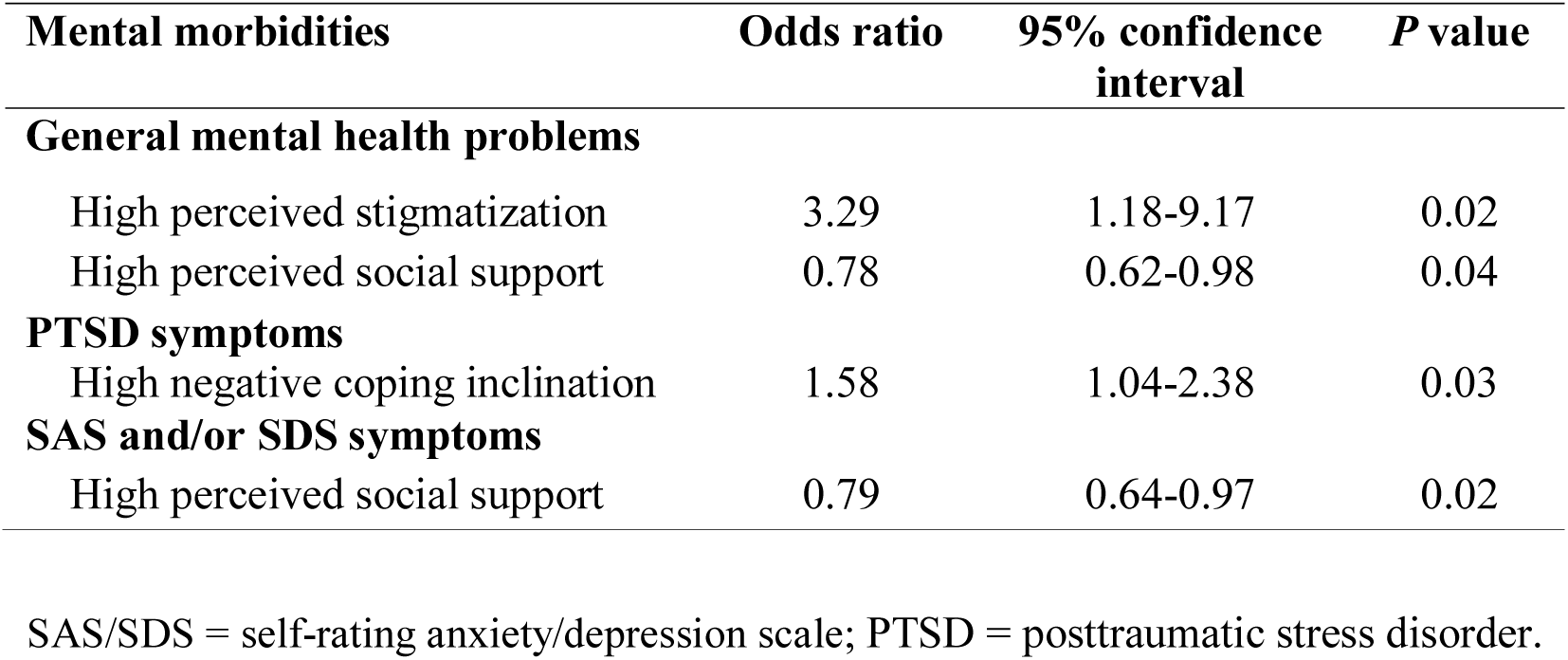
Logistic regression with variables predicting main outcomes of mental morbidities.

| **Mental morbidities** | **Odds ratio** | **95% confidence interval** | ***P* value** |
| --- | --- | --- | --- |
| **General mental health problems** |  |  |  |
| High perceived stigmatization | 3.29 | 1.18-9.17 | 0.02 |
| High perceived social support | 0.78 | 0.62-0.98 | 0.04 |
| **PTSD symptoms** |  |  |  |
| High negative coping inclination | 1.58 | 1.04-2.38 | 0.03 |
| **SAS and/or SDS symptoms** |  |  |  |
| High perceived social support | 0.79 | 0.64-0.97 | 0.02 |

SAS/SDS = self-rating anxiety/depression scale; PTSD = posttraumatic stress disorder.

### PTSD symptoms in COVID-19 patients

5 out of 41 patients (12.2%) had PTSD symptoms. Compared to patients without PTSD symptoms, those with PTSD symptoms had, on average, a lower objective social support score, but higher negative coping style scores and higher perceived social stigmatization (**Table S1**). Results from logistic regression analysis indicated that having a high negative coping style score (OR, 1.58; 95% CI, 1.04–2.38; *P* = 0.03) was associated with a greater risk of PTSD symptoms in patients (**Table 3**).

### Anxiety and/or depression symptoms in COVID-19 patients

Among the 41 patients, 11 (26.8%) had anxiety and/or depression symptoms (5 had both anxiety and depressive symptoms, 5 had only depressive symptoms, and 1 had only anxiety symptoms). Compared to patients without anxiety and/or depression symptoms, those with anxiety and/or depression symptoms had lower objective and perceived social support scores (**Table S2**). Results of logistic regression analyses indicated that high perceived support score (OR, 0.79; 95% CI, 0.64-0.97; P = 0.02) was associated with a lower risk of anxiety and/or depression symptoms in patients (**Table 3**).

### Chronic fatigue in COVID-19 patients

22 (53.6%) patients reported chronic fatigue problems. Relative to patients without chronic fatigue problems, those with fatigue problems had a lower perceived social support score (**Table S3**). However, logistic regression analysis did not detect any association between clinical, psychological measures and the risk of fatigue problems in patients.

## DISCUSSION

In this study, we investigated psychological morbidities and fatigue in patients with confirmed COVID-19 during the outbreak. Results demonstrated that the rates of general mental health problems, psychological morbidities, and chronic fatigue are very common among COVID-19 patients. The mental health problems in COVID-19 patients were alarming. Specifically, we found that being stigmatized and negative coping inclination are the main risk factors, while high perceived social support is the main protective factor for COVID-19 patients’ mental health. No risk or protective factors were found concerning fatigue; no relationships were detected between age, sex, educational level, marital status, pre-existing disabilities, clinical symptoms, current severity of pneumonia and mental health in patients participating in this study. The findings in this study shed light on the need for proactive social support and care for the mental health in COVID-19 patients during the epidemic.

Presence of psychological morbidities and chronic fatigue are common in COVID-19 patients who participated in this study. COVID-19 was not merely an episode of illness for the infected patients, but a life-changing disastrous experience, which not only impairs physical well-being but also their mental health. Rates of psychological morbidities found in this study were higher than those reported among the nationwide general population in a China Mental Health Survey^23^, in which anxiety (lifetime prevalence: 7.6%) was the most common mental health condition. However, the rate presented here was consistent with extant literature on prior public health emergencies such as SARS, which reported rates of anxiety and/or depression between 15.6%^24^ to 35%^25^; chronic fatigue between 17.7%^26^ to 40.3%^10^; and PTSD between 6%^27^ to 42.1%^28^ in SARS patients during and/or after the disease outbreak. Impaired general mental health was also very common in SARS^29^ and MERS^6^ survivors compared to non-SARS/MERS survivors (albeit no exact incidence rate had been reported). Notably, prior studies demonstrated that mental health problems in SARS survivors such as PTSD and chronic fatigue - could be detected over the long term, according to 4-year followups.^10,28^ Thus, we forecast that the COVID-19 outbreak would also likely result in persistent psychological impact in infected patients. These results highlight the need to enhance the preparedness and competence of health care professionals to detect and manage the psychological sequelae of the currently emerging COVID-19 epidemic.

### Risk factors for psychological morbidities in COVID-19 patients

In the present study, being stigmatized was found to be a contributing factor for impaired general mental health, and a negative coping inclination was a contributing factor for PTSD symptoms. Disease-associated stigma is complex, and may be present in both the acute and recovery phases of the disease. In the acute phase, fear of contagion, unclear pathologic characteristics, and being subjected to quarantine measures^30^ could cause disproportionate and undesirable labeling of the patients and even their families. In the recovery phase, the residual physical symptoms and chronic fatigue are often viewed as dubious and controversial. Siu et al. showed that SARS-related stigmatization in SARS survivors persisted in a 16-month observation^31^. In a follow-up study, perceived stigmatization of SARS survivors predicted a greater risk of psychiatric morbidity in the long term^10^. Therefore, our finding of stigmatization as a risk factor for mental health problems in COVID-19 patients aligns with prior findings in SARS patients.

Negative coping styles were related to post-traumatic symptoms in the first-time mothers^32^, and related to greater anxiety and depression in accident and emergency senior house officers^33^. Findings from all these studies collectively suggest that education for patients in self-coping strategies may help to mitigate their mental morbidities.

### Protective factor of psychological morbidities in COVID-19 patients

In this study, high perceived social support is a protective factor for general mental health, and against anxiety and/or depression symptoms in patients. Unlike objective support, perceived support pertains to the feeling of being supported, and has been thought to be a more useful social support subscale^34,35^. In a prior study, perceived support was associated with the recovery from prior PTSD, but objective support had no such association^35^. Our finding is also consistent with previous findings regarding the role of perceived social support during the SARS epidemic. Low emotional support was associated with anxiety and depression symptoms in SARS survivors^13^. Also, health care workers who experienced psychiatric symptoms during the SARS epidemic were more likely to report that they did not receive enough support from their supervisor or head of department^9^. This shows that the strain on mental health during an epidemic affects health care providers and patients alike. Our findings suggest the need for healthcare institutions to provide proactive psychological support for COVID-19 patients to enhance their resilience to mental morbidities.

### Effects of Physical states on psychological morbidities in COVID-19 patients

To the best of our knowledge, no study till now has directly investigated the effects of physical states on psychological morbidities in COVID-19 patients. In this study, we used several physical indices – physical comorbidity, clinical symptoms, and severity of pneumonia – to examine their effect on mental health in COVID-19 patients. Unfortunately, the result showed that the mental health in patients was not influenced by their physical states, which was inconsistent with our original hypothesis. This negative result suggests that the mental health in COVID-19 patients was mainly affected by the psychological rather than the biological situation. Future studies are warranted to verify this finding.

### Limitations

Our study had several limitations. First, the results in this study are limited to a small sample size of patients with non-severe disease type. The association between disease severity and psychological morbidities and fatigue, although not found in this study, should be further assessed by recruiting more patients with severe disease. Second, we originally designed this study to investigate both the prevalence and associated biopsychosocial risk factors (including clinical symptoms and disease severity parameters) for mental health problems. Suspected cases of COVID-19 who were quarantined in homes, hotels, and hospitals were not included in this study. However, these suspected cases may also have psychological morbidities especially if they have been facing considerable mental stress – fear of contagion, feeling frightened, and helplessness^3,30^. The psychosocial impact of the COVID-19 outbreak on suspected cases needs to be clarified in future studies. Third, the questionnaires used in this study are brief and self-reported. As patients are easily tired and are under treatment in isolation wards^8^, face-to-face psychiatric assessment was not conducted in the present study. A formal post-discharge evaluation of psychometric properties via psychological experts should be conducted for these patients.

## CONCLUSION

In conclusion, during the disease outbreak, psychological morbidities and chronic fatigue are common in COVID-19 patients. Being stigmatized and negative coping strategies are the main risk factors, while high perceived social support is the main protective factor of mental health in patients. These findings shed light on the need for healthcare institutions to be aware of mental health morbidities in patients during the COVID-19 epidemic.

## Data Availability

We state that Dr. Rongfeng Qi (the first author) takes full responsibility for the data, analyses, interpretations, and the research, he will also represent us for contacting with your journal. Prof.Guang Ming Lu has full access to all of the data, and has the right to publish any and all data separate and apart from any sponsor.

## ACKNOWLEDGMENTS

This work was funded by the grants from the National Nature Science Foundation of China [Nos. 81671672, 81301209 to R.Q.]; the Jiangsu Provincial Medical Youth Talent [Nos. QNRC2016888 to R.Q.]; the 333 high-level talents training project of Jiangsu province [No. (2018) Ⅲ-2375 to R.Q.]; the Chinese Key Grant [No. BWS11J063 to G.M.L.]; and NIH [No. U54 EB020403 to PMT].

